# Evidence of SARS-CoV-2 JN.2.5 variant circulation in Rio de Janeiro, Brazil

**DOI:** 10.1101/2024.04.25.24306286

**Authors:** Mariane Talon de Menezes, Camila de Almeida Velozo, Filipe Romero Rebello Moreira, Diana Mariani, Érica Ramos dos Santos Nascimento, Cássia Cristina Alves Gonçalves, Lídia Theodoro Boullosa, Thais Félix Cordeiro, Gleidson Silva Oliveira, Maria Cecília da Cunha Carneiro, Cinthia Francisca Valdez, Natacha Cunha de Araujo Faria, Bianca Ortiz da Silva, Rafael Mello Galliez, Átila Duque Rossi, Carolina Moreira Voloch, Terezinha Marta Pereira Pinto Castiñeiras, Amilcar Tanuri

## Abstract

New SARS-CoV-2 variants are continually emerging and have the potential to present challenges to public health responses worldwide. This report discusses the early detection of the lineage JN.2.5 (alias of B.1.1.529.2.86.1.2.5) in Rio de Janeiro, Brazil. The variant was identified in early 2024 through viral whole-genome sequencing and phylogenetic analysis, revealing that circulating strains present genetic similarities to sequences reported in Canada, Poland, Belgium, and England. While the circulation of diverse Omicron sub lineages has significantly impacted COVID-19 epidemiology in Brazil, the consequences of the circulation of the novel variant are uncertain. Considering its immunological escape capabilities and epidemiological behavior observed elsewhere, close monitoring of this variant epidemiology is recommended.

## Introduction

New Severe Acute Respiratory Syndrome Coronavirus 2 (SARS-CoV-2) variants continue to emerge and have the potential to present challenges to global public health. Since its emergence in late 2021, several lineages derived from the Omicron variant of concern have spread globally, dominating epidemiological scenarios and alternating in frequency through antigenic drift (Carabelli et al. 2023, Yang et al. 2024). More recently, the Omicron lineage BA.2.86.1 has given rise to several sub lineages, including JN.1 and JN.2.5, both of which possess the S:L455S substitution in the spike protein (Roemer 2024). Studies have shown that this substitution provides the JN.1 variant with enhanced immune evasion capabilities (Yang et al. 2024). JN.1 has also shown increased transmissibility, becoming the predominant variant worldwide in the early months of 2024 and correlating with a rise in COVID-19 cases (Kaku et al. 2024). Currently, JN.2.5 is most prevalent in Canada, accounting for approximately 92% of all JN.2.5 deposited genomes (Pangolin 2024). In January 2024, the JN.2.5 variant was first detected in Brazil in Mato Grosso do Sul state, Central-West region of the country (Brasil 2024b).

Here, we present a SARS-CoV-2 genomic surveillance study involving a cohort of 47 symptomatic individuals attending the Núcleo de Enfrentamento e Estudos de Doenças Infecciosas Emergentes e Reemergentes (NEEDIER) at Universidade Federal do Rio de Janeiro (Rio de Janeiro, Brazil), between November 2023 and February 2024. All individuals were confirmed positive for SARS-CoV-2 by RT-qPCR (CT < 26) of nasopharyngeal swab samples. Total viral RNA was extracted from swabs using the KingFisher Flex System (ThermoFisher Scientific, Waltham, USA) following the manufacturer’s instructions. cDNA synthesis was then performed using the LunaScript RT SuperMix Kit (New England Biolabs, Massachusetts, USA). The ARTIC Network SARS-CoV-2 version v4.1 primer scheme and Q5 High-Fidelity DNA polymerase were employed for SARS-CoV-2 whole-genome multiplex PCR amplification. Following PCR amplification, the products were used to construct a library using the Rapid Barcoding kit (ONT SQK-RBK114), following the manufacturer’s instructions. Sequencing was conducted using an R9.4.1 MinION flow cell (Oxford Nanopore Technologies, Oxford, UK). Consensus genome sequences, inferred with the ARTIC pipeline, were submitted to the Gisaid database (ref) (EPI_SET_240311rg). Metadata and assembly statistics are available in Supplementary Table 1.

Altogether, 47 SARS-CoV-2 genome sequences were classified with NextClade v3.3.1 (Aksamentov et al. 2021) and Pangolin v4.3.1 (O’Toole et al. 2021). A total of 11 Pango lineages were identified: GK.1.10 (1), GK.1.2.1 (2), GK.1.8 (1), JD.1.1 (5), JN.1 (27), JN.1.1 (1), JN.1.16 (2), JN.1.18 (2), JN.2.5 (3), XBB.1.16.6 (1), XDR (2). Temporal lineage frequency analysis of these sequences revealed the predominance of the JN.1 variant in Rio de Janeiro during the early months of 2024. This analysis is consistent with additional data available on EpiCoV (**Supplementary Figure 1**). Moreover, the analysis identified the introduction of the JN.2.5 variant in the city in February (**Figure 1**).

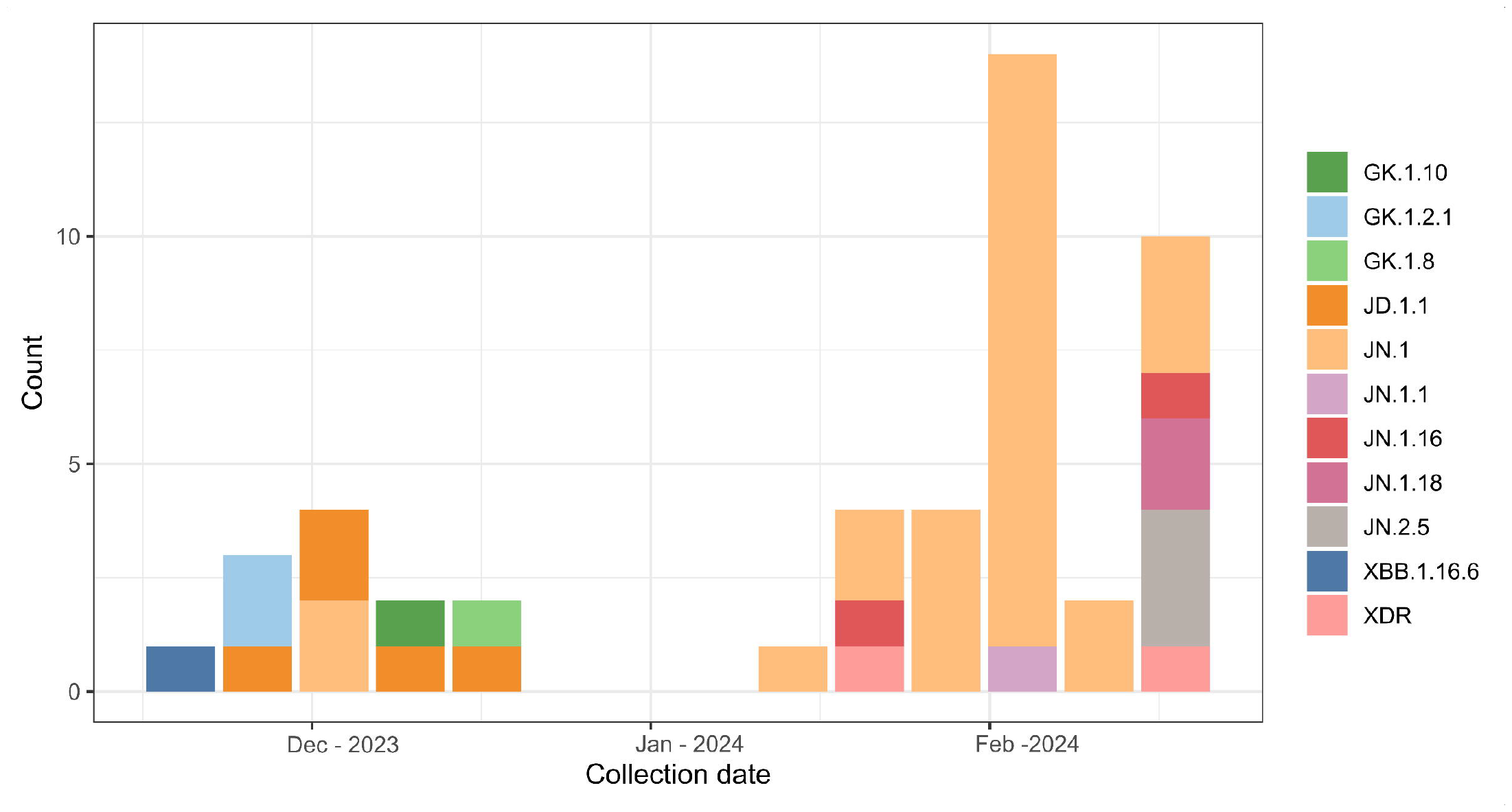

To further contextualize the novel sequences, a comprehensive phylogenetic analysis was performed. A reference dataset was constructed using data available on GISAID EpiCoV; briefly, we used the Audacity instant app to retrieve the 20 sequences most similar to each of the sequences generated in this study. Additionally, we included all Brazilian sequences available between November 2023 and February 2024. Sequences were aligned using the software minimap2 (Li 2018) and gofasta (Jackson 2022), and a maximum likelihood phylogenetic tree was inferred with IQ-TREE (Minh et al. 2020) under the best-fit model, as identified by ModelFinder (Kalyaanamoorthy et al. 2017). Shimoidara-Hasegawa-like approximate likelihood ratio test (SH-aLRT) was used to calculate nodes’ statistical support (Guindon et al. 2010).

The analysis revealed two well-supported clades, confirming previous classifications (**Figure 2**). One clade consisted of sequences that evolved from the BA.2.86.1 lineage ancestor, including JN.1, JN.2.5, and XDR (a JD.1.1.1 and JN.1.1 recombinant sub lineage). The other clade (XBB*) exhibited a different Omicron diversity, containing the XBB, GK, and JD variants. It is noteworthy that all variants in the XBB descendants clade were found to be circulating in the city only before January 2024, indicating a phylogenetic shift in this period, possibly driven by antigenic drift (Yewdell 2021).

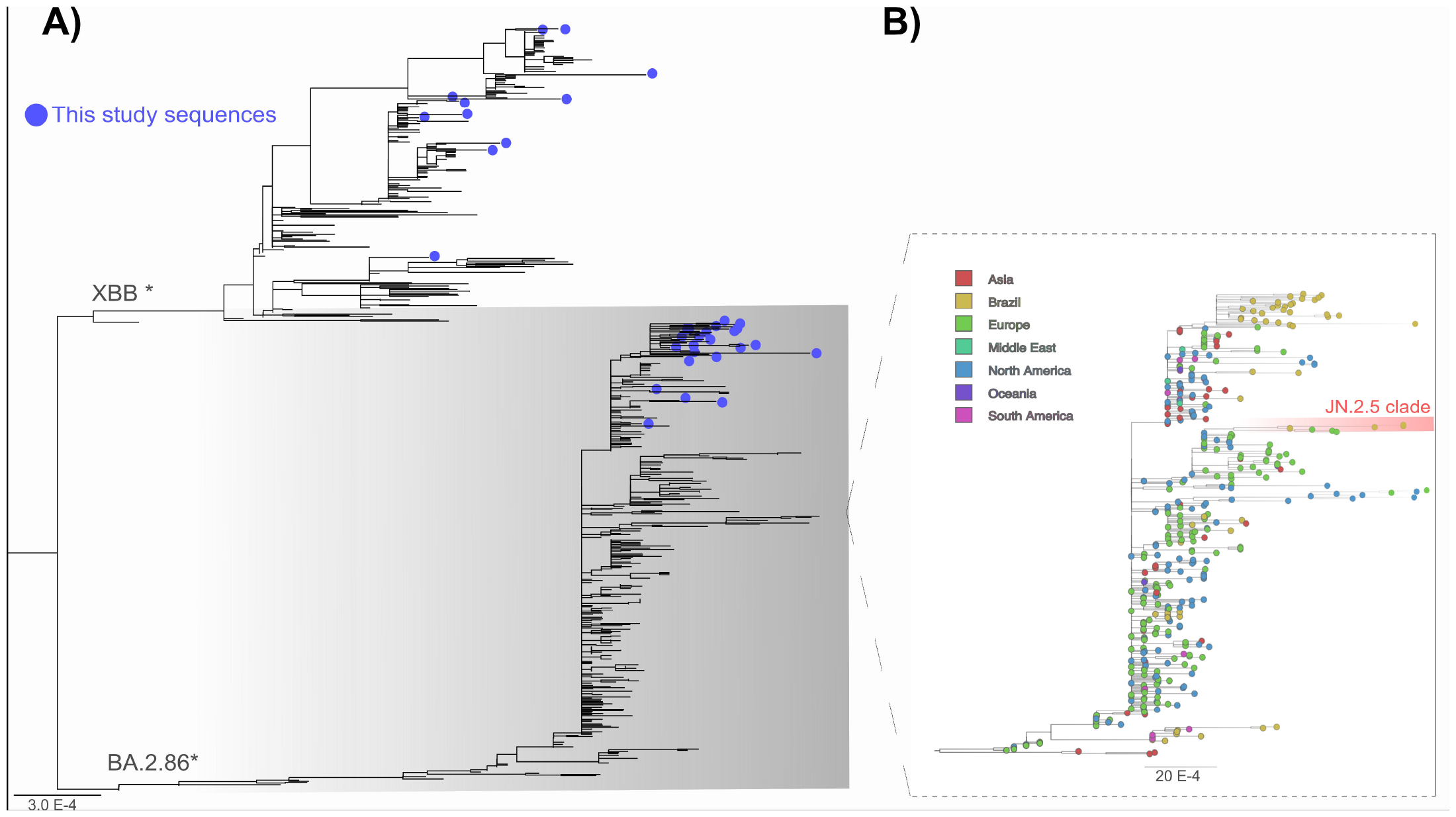

The introduction of JN variants between December 2023 and January 2024 may be related to holidays and vacation trips in the country. Indeed, Brazilian health authorities reported an increase in COVID-19 cases after this period (Brasil 2024a). One limitation of this study is the lack of data in December 2023, missing the exact moment when the variant profile shift occurred. This limitation is also present in the GISAID database and represents a bias in genomic surveillance activities in the country during holiday seasons, as observed elsewhere (**Supplementary Figure 1)** (Chen et al. 2020).

Given that Rio de Janeiro is a major hub of tourism and business - one of the most visited cities in the Southern Hemisphere - our results emphasize the role of global mobility in sustaining international chains of transmission, as previously noticed (Tegally et al. 2023). Regarding the variant JN.2.5, our analysis suggests it harbors a high degree of genetic similarity to sequences previously reported in Canada, Poland, Belgium, and England. Additionally, a JN.1 clade generated by sequences from our study underlines the occurrence of local transmission chains for this variant in Rio de Janeiro. Further analysis should explore the epidemiological, serological, and virological properties of lineage JN.2.5, focusing on clarifying its potential risk for global public health.

## Acknowledgements

We thank all authors who submitted their data to GISAID EpiCoV. A full acknowledgment table can be found in https://doi.org/10.55876/gis8.240419fw (EPI_SET_240419fw).

## Institutional Review Board Statement

The study was conducted according to the guidelines of the Declaration of Helsinki, and approved by the Ethics Committee of Hospital Universitário Clementino Fraga Filho (protocol numbers 30161620.0.1001.5257).

## Data Availability Statement

All generated genome sequences have been deposited on GISAID EpiCoV (EPI_SET_240311rg).

## Sponsorships

This work was supported in part by grants from Fundação Carlos Chagas Filho de Amparo à Pesquisa do Estado do Rio de Janeiro/FAPERJ (E−26/210.785/2021) and from Instituto Todos pela Saúde/ITpS (C1114/2021). These sponsors had no role in the study design, execution, analysis, interpretation of data, or decision to present the results.

