## Supplementary figures and images for "Evidence of SARS-CoV-2 JN.2.5 variant circulation in Rio de Janeiro, Brazil"

### Supplementary Figure 1

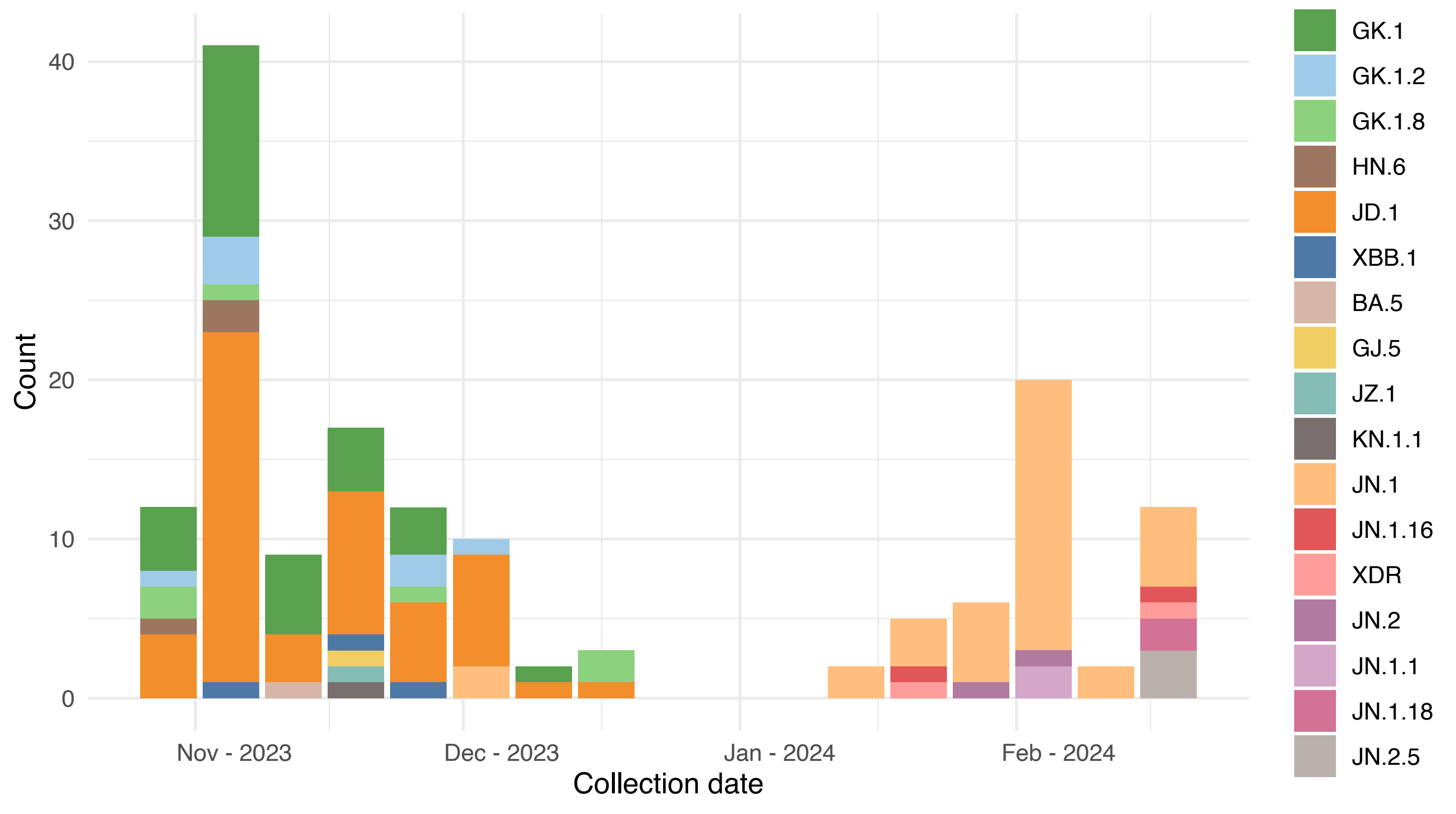
